# Cross-cultural adaptation and psychometric validation of the ISBAR Structured Handover Observation Tool in ICU-to-ward patient transfer

**DOI:** 10.64898/2026.04.10.26350669

**Authors:** Na Ni, Yuting Wang, Qi Wang, Jiajia Ding, Tong Liu, Bixia Zhao

## Abstract

The ISBAR framework is used to standardize clinical handovers and enhance patient safety. Observational tools based on ISBAR have been developed to assess the completeness of information transfer. However, these instruments have primarily been developed in non-Chinese contexts, and validated Chinese-language observational tools suitable for clinical practice remain limited. In this study, a cross-cultural adaptation and psychometric validation of the ISBAR Structured Handover Observation Tool was conducted, examining its reliability and discriminant validity in Chinese clinical settings. The study was conducted in two phases: cross-cultural adaptation and psychometric evaluation in real-world clinical settings. Content validity was assessed using the Content Validity Index (CVI), and inter-rater reliability was evaluated using the Intraclass Correlation Coefficient (ICC) based on a two-way mixed-effects model with absolute agreement. Discriminant validity was examined using the Mann–Whitney U test to compare scores across nurses with varying levels of clinical experience. A total of 233 handover cases involving patient transfers from the intensive care unit (ICU) to general wards were collected, involving 84 nurses. The scale demonstrated good content validity, with item-level content validity indices (CVI) ranging from 0.88 to 1.00 and a scale-level CVI/Ave of 0.98. The inter-rater reliability, assessed using fifty randomly selected cases, was high, with an intraclass correlation coefficient (ICC) of 0.885 for single-rater assessments and 0.939 for average-rater assessments. Discriminant validity analysis showed that nurses with more clinical experience had significantly higher total scores than those with less experience (Z = −4.772, p < 0.001). The Chinese version of the ISBAR Structured Handover Observation Tool demonstrates good content validity, high inter-rater reliability, and acceptable discriminant validity. This tool provides a standardized and practical method for assessing the completeness of information transfer and is expected to support quality improvement in patient handover from the ICU to general wards in Chinese clinical settings.

## Introduction

An effective clinical handover is a critical component of patient safety, ensuring continuity of care and the accurate transmission of clinical information [1–3]. Communication errors during handoffs have been identified as a major cause of adverse events in healthcare settings [4]. This is especially true during high-risk transfers, such as when moving patients from the intensive care unit (ICU) to a general ward [5]. The Joint Commission reports that communication breakdowns are the primary root cause of sentinel events, accounting for a significant proportion of serious medical errors [6]. These referral processes involve complex clinical situations, a high volume of information, and changes in nursing responsibilities, which increase the risk of information loss and communication errors [7].

To improve the quality and consistency of clinical communication, structured communication tools such as ISBAR (Identity, Situation, Background, Assessment, Recommendation) have been widely recommended [6,8]. ISBAR provides a structured framework for organizing and conveying critical patient information [9–12], and previous studies have confirmed its effectiveness in improving communication quality and reducing adverse events.

However, the effectiveness of structured communication tools depends on whether they are applied consistently and accurately in clinical practice. Evidence suggests that without systematic evaluation and feedback, adherence to structured communication protocols may decline over time [13]. Therefore, objective and reliable methods are needed to assess whether ISBAR is being applied correctly during clinical handoffs.

To address this need, Tangvik et al. developed the ISBAR Structured Handover Observation Tool, which was specifically designed to assess the use of ISBAR during patient transfers from intensive care units to general wards [1]. Unlike general communication assessment tools, this tool focuses on whether predefined key information elements are reported, thereby enabling an objective evaluation of adherence to structured communication protocols [1,7,14]. However, evaluating only the completeness of information may not fully reflect the quality of the handover; a complete handover is not necessarily an effective one, as the latter also involves dimensions such as clarity of explanation and interactive confirmation. There is still ongoing methodological debate regarding how to balance the objectivity and comprehensiveness of such evaluations.Existing assessment tools have been primarily developed and validated in non-Chinese contexts, with no validated Chinese versions currently available [15,16]. This gap limits the ability to objectively assess the aforementioned handover processes in clinical settings in China. Furthermore, the applicability of these tools in actual clinical practice—particularly during the transfer from the ICU to general wards—has not been fully explored. Given the complexity and diversity of such transfer processes, there is a need for a culturally adapted and psychometrically validated observational tool to support standardized assessment and quality improvement.

Therefore, this study aims to conduct a cross-cultural adaptation and psychometric validation of the ISBAR Structured Handover Observation Tool and to examine its reliability and validity in assessing the completeness of information transfer during patient handover from the ICU to general medical wards in a Chinese clinical setting. This study successfully achieved the aforementioned objectives and reported the reliability and validity results of the Chinese version of the tool in assessing the completeness of information transfer when patients are transferred from the ICU to general medical wards in a Chinese clinical setting.

## Method

### Study design

This methodological study was conducted in two phases: the cross-cultural adaptation and psychometric validation of the ISBAR Structured Handover Observation Tool.

### Study setting

This study was conducted in the ICUs and general medical wards of two tertiary hospitals. Data were collected during the routine clinical handover process when patients were transferred from the ICU to the general medical ward, providing an accurate reflection of clinical practice.

### Participants and sampling

The study subjects were nurses involved in the handover process from the ICU to general medical wards.

A total of 233 handover cases from the ICU to general wards were collected from two tertiary hospitals, involving 84 nurses.

To conduct an inter-rater reliability analysis, 52 handover cases were selected as a subsample; two of these were excluded due to incomplete or interrupted handover processes. Ultimately, 50 eligible cases were included in the ICC analysis.

The inclusion criteria for the study were as follows: ① Nurses must hold a valid nursing license and must have worked in the department for at least six months; ② The patient was transferred from the ICU to a general medical ward; ③ The nurses should have participated in routine handover tasks during the study period and voluntarily agreed to participate in this study; ④ The handover process was completed, allowing for full observation and documentation. The exclusion criteria were as follows: ① Intern nurses, visiting nurses, or those who had worked in the department for less than six months; ② The handover process was incomplete or interrupted; ③ The patient was transferred out of the department to a non-ICU unit or transferred into a department other than a general medical ward.

To test whether the ISBAR observation tool could effectively distinguish between groups of nurses with varying levels of clinical experience, the nurses were divided into two groups based on their years of service: the junior group (years of service < 5 years, n = 44) and the senior group (years of service ≥ 5 years, n = 40). In domestic nursing research literature, junior nurses are commonly defined as those with five years or less of work experience [17], and previous studies have confirmed that the number of years of experience is significantly associated with communication quality [18]. Therefore, this study is also suitable for testing known-group validity. The Mann-Whitney U test [19] was used to compare the differences in the total scores on the ISBAR observation tool between the two groups of nurses, with the expectation that the senior group’s scores would be significantly higher than those of the junior group. The baseline characteristics of the two groups, including age, professional title, and educational background, will be presented in detail in the results section.

### Instrument

The ISBAR Structured Handover Observation Tool, originally developed by Tangvik et al. [1], is an observation checklist designed to assess the completeness of information transfer during patient handoffs.

Based on the ISBAR framework, the tool comprises five dimensions: Identification, Situation, Background, Assessment, and Recommendation. It consists of 26 items, each representing a key piece of information that should be conveyed during the handoff.

Each item is scored using a binary scale (0 = missing, 1 = present) based on whether the relevant information was reported. The total score is calculated by summing the scores of all items; a higher score indicates greater completeness of information transfer.

### Cross-cultural adaptation

The cross-cultural adaptation of the ISBAR Structured Handover Observation Tool was conducted in accordance with the established guidelines proposed by Beaton et al. [20]. Formal permission from the original authors was obtained in advance of the adaptation process.

The adaptation procedure consisted of five steps to ensure semantic, conceptual, and cultural equivalence between the original and Chinese versions:

1. Forward translation: Two bilingual nursing researchers independently translated the original instrument into Chinese. This process yielded two initial Chinese versions (T1 and T2).
2. Consolidation: The two translations were compared and consolidated into a preliminary Chinese version (V1). Discussions were held to reach a consensus and resolve discrepancies.
3. Back-translation: Two translators, unaware of the original scale’s content, independently back-translated the consolidated version (V1) into English. This generated two back-translated versions (RT1 and RT2), which were subsequently integrated into a single version.
4. Expert Panel Review: A seven-member expert panel was convened, comprising four ICU specialists, two bilingual translators, and one researcher. The panel assessed the conceptual equivalence of the back-translated version with the original scale and, through discussion and consensus, produced the pre-final Chinese version (V2).
5. Pilot Testing: In a clinical setting, the pre-final version was pilot-tested during 15 nurse-led patient handover processes to assess its clarity, feasibility, and comprehensibility. Each handover was independently evaluated by two trained assessors. Based on their feedback and observations, the text was fine-tuned to produce the final Chinese version. Detailed records of the translation and adaptation process are available in Supplementary Data S3.

### Data collection

In routine clinical settings, handover processes during patient transfers from the ICU to general wards were observed. Each handover was independently assessed by two trained evaluators using the ISBAR Structured Handover Observation Tool. To ensure objectivity, the evaluators did not participate in patient care or the handover process.

Both evaluators were experienced clinical nurses with over 10 years of experience in either the ICU or general wards. Prior to data collection, they received approximately two hours of standardized training. The training included an introduction to the ISBAR framework, detailed explanations of each item, and clarification of the scoring criteria.

To ensure consistency, the evaluators jointly assessed three pilot handover cases and discussed any discrepancies to reach consensus on the scoring criteria. A preliminary inter-rater reliability assessment based on these pilot cases demonstrated good agreement (ICC > 0.80). During the formal data collection phase, each handover event was independently assessed by two raters; regular discussions were held to resolve discrepancies and maintain consistency.

All handover processes were video-recorded, and observers independently scored the video footage afterward to ensure that the observation activities did not interfere with routine clinical practice.

### Data analysis

All statistical analyses were performed using SPSS Version 26.0 (IBM, Armonk, NY, USA).

Content validity was assessed using the content validity index (CVI). Inter-rater reliability was assessed using the ICC, based on a two-factor mixed-effects model of absolute agreement [21]. The Mann-Whitney U test was used to compare scores among nurses with different levels of clinical experience to assess discriminant validity [19].

A p-value < 0.05 was considered statistically significant.

The sample size was deemed adequate for reliability analysis, as previous methodological studies have indicated that at least 30 observations are sufficient to obtain stable estimates of inter-rater agreement [22,23]. The raw data are available in Supplementary Data S1.

### Ethical considerations

This study has been approved by the Medical Ethics Committee of the University of South China (Approval No.: 2025-0148) and was conducted in accordance with the Declaration of Helsinki.

Data were collected from October 1, 2025, to February 15, 2026. Prior to data collection, all eligible nurses were provided with an information sheet explaining the study purpose, procedures, risks, benefits, confidentiality, and voluntary nature of participation. Written informed consent was obtained from each nurse by having them sign a unified consent form. All data were anonymized to ensure confidentiality.

## Results

### Cross-cultural adaptation

The translation and back-translation processes were completed smoothly without encountering any significant difficulties, and the overall structure of the original scale was preserved.

Based on the expert panel’s assessment, minor adjustments were made to the scale to enhance the semantic clarity and cultural relevance. These revisions primarily involved optimizing the wording and modifying the item descriptions to better reflect local clinical practice. Detailed records of the translation, adaptation, and expert consultation can be found in Supplementary Data Sets S2 and S3.

Based on expert consensus, one item related to communication regarding community services was deleted. Most experts noted that, in the Chinese clinical context, community-based healthcare services have not yet been widely implemented, which limits the applicability of this item in routine handoffs. Furthermore, factors such as nursing needs and social support were considered to be adequately covered by other items, potentially leading to content duplication. Therefore, the expert panel ultimately agreed to delete this item to enhance the scale’s cultural relevance and overall feasibility.

### Content validity

In the first round of expert consultation, the item-level content validity indices (I-CVI) ranged from 0.67 to 1.00, with S-CVI/UA at 0.62 and S-CVI/Ave at 0.96.

Following revisions, the results from the second round improved, with I-CVI ranging from 0.88 to 1.00, S-CVI/UA at 0.76, and S-CVI/Ave at 0.98 (Table 2). Detailed records of the expert consultation are available in Supplementary Data S2.

These results indicate that the content validity of the scale is at an acceptable to excellent level.

### Reliability

#### Characteristics of case handover

A total of 50 eligible transfer cases were included in the inter-rater reliability analysis. The mean age of the patients was 68.6 years (standard deviation = 10.43), and the mean length of stay in the ICU was 3.14 days (standard deviation = 1.58). The mean total scores assigned by the two raters were comparable (19.50 ± 3.65 vs. 19.08 ± 3.52), indicating good inter-rater agreement. Patients were transferred to multiple departments, with the majority admitted to the Department of Gastroenterology (32.0%), followed by the Department of Neurology (20.0%), the Department of Cardiology (18.0%), and the Department of Pulmonology (16.0%).

#### Inter-rater reliability

The ISBAR observation tool demonstrated good inter-rater reliability. The single-measurement ICC for the total score was 0.885 (95% CI: 0.795–0.935), and the average-measurement ICC was 0.939 (95% CI: 0.886–0.966). The single-measurement ICC values for each dimension ranged from 0.789 to 0.941, with the reliability for all dimensions ranging from good to excellent (Table 1).

**Table 1.** Reliability of the ISBAR Structured Handover Observation Tool.

| Dimension | ICC (single) | 95% CI |
| --- | --- | --- |
| Total | 0.885 | 0.795–0.935 |
| Identification | 0.941 | 0.898–0.966 |
| Situation | 0.913 | 0.853–0.950 |
| Background | 0.917 | 0.858–0.952 |
| Assessment | 0.789 | 0.634–0.880 |
| Recommendation | 0.799 | 0.671–0.880 |

*Note: ICC, intraclass correlation coefficient; CI, confidence interval. ICC values $\geq 0.75$ indicate good reliability, and $\geq 0.90$ indicate excellent reliability.*

### Validity

#### Baseline characteristics of nurses

This study included a total of 84 nurses, with 44 in the junior group (i.e., less than five years of experience) and 40 in the senior group (i.e., five or more years of experience). The distribution of age, gender, professional title, educational background, and the number of handover sessions attended in the past month for both groups is shown in Table 2.

**Table 2.**
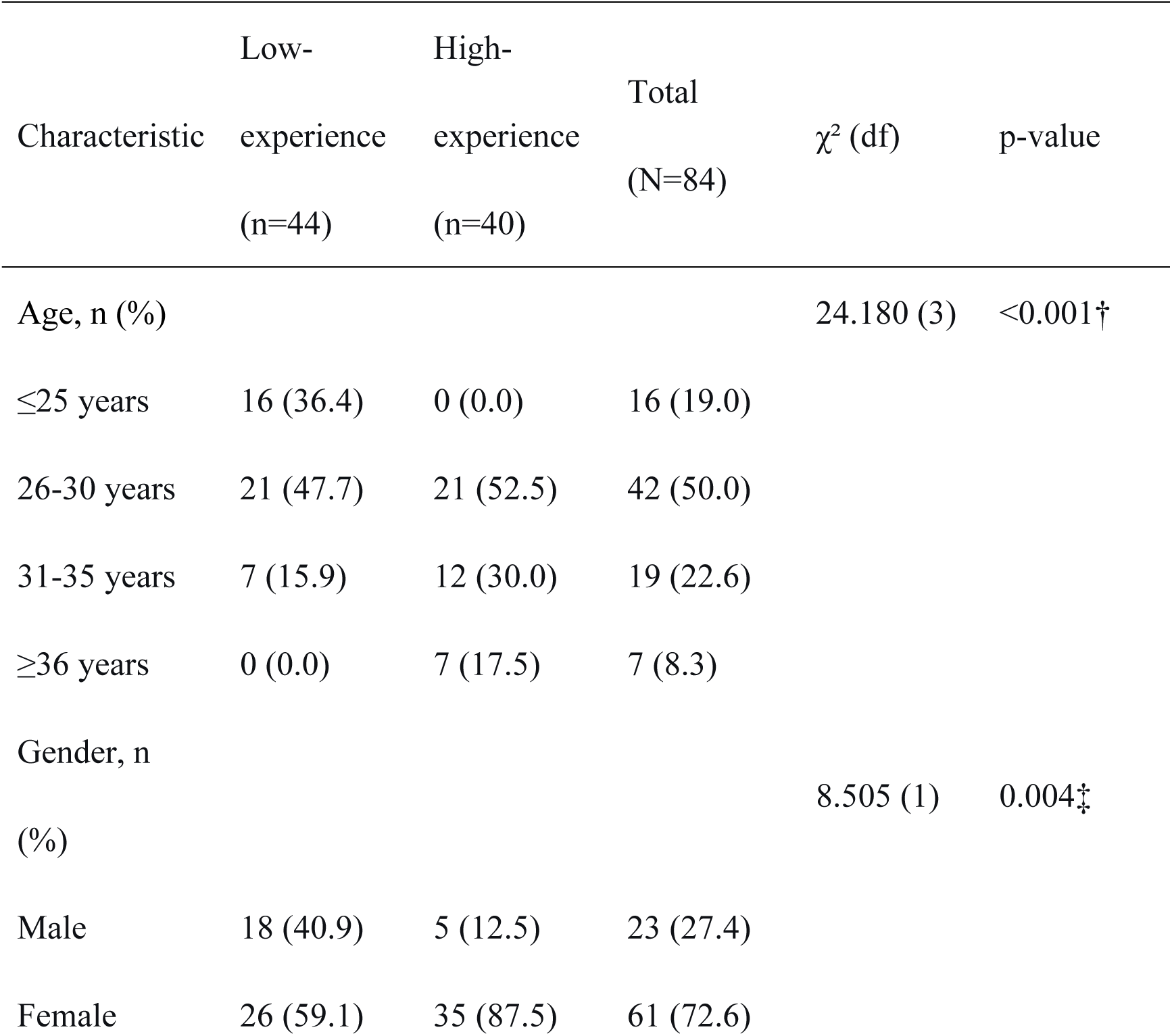
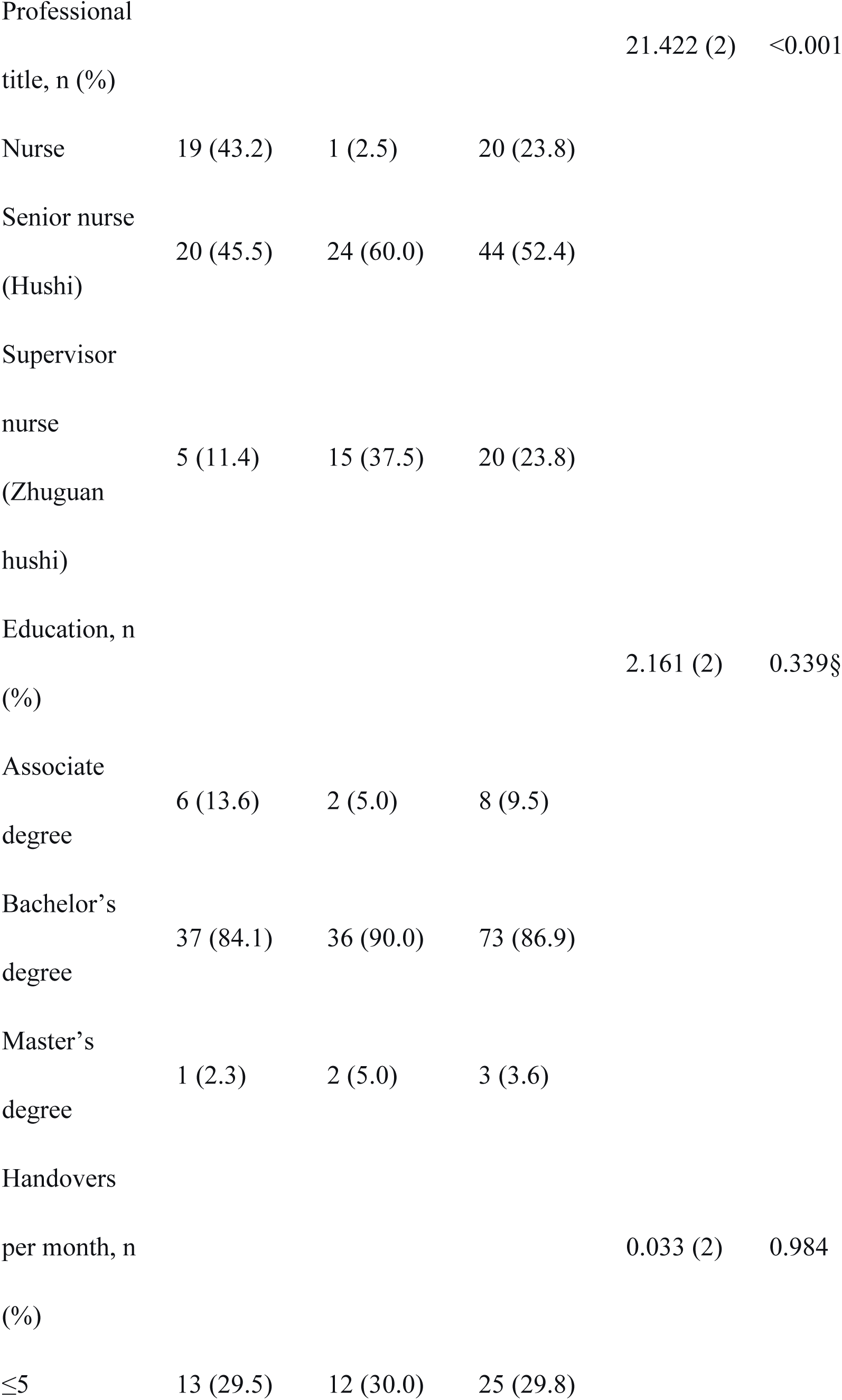
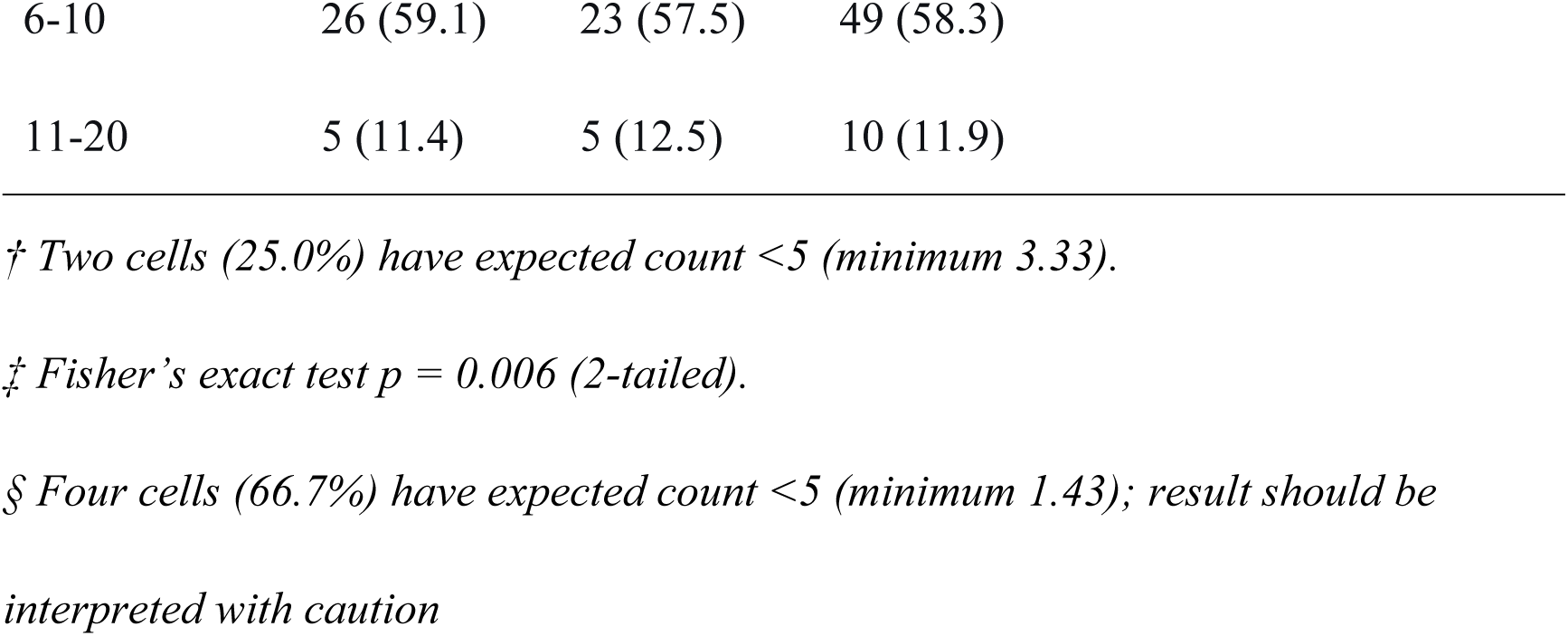
Baseline characteristics of nurses by experience group.

| Characteristic | Low-<br>experience<br>(n=44) | High-<br>experience<br>(n=40) | Total<br>(N=84) | $\chi^2$ (df) | p-value |
| --- | --- | --- | --- | --- | --- |
| Age, n (%) |  |  |  | 24.180 (3) | <0.001† |
| ≤25 years | 16 (36.4) | 0 (0.0) | 16 (19.0) |  |  |
| 26-30 years | 21 (47.7) | 21 (52.5) | 42 (50.0) |  |  |
| 31-35 years | 7 (15.9) | 12 (30.0) | 19 (22.6) |  |  |
| ≥36 years | 0 (0.0) | 7 (17.5) | 7 (8.3) |  |  |
| Gender, n (%) |  |  |  | 8.505 (1) | 0.004‡ |
| Male | 18 (40.9) | 5 (12.5) | 23 (27.4) |  |  |
| Female | 26 (59.1) | 35 (87.5) | 61 (72.6) |  |  |
| Professional<br>title, n (%) |  |  |  | 21.422 (2) | <0.001 |
| Nurse | 19 (43.2) | 1 (2.5) | 20 (23.8) |  |  |
| Senior nurse<br>(Hushi) | 20 (45.5) | 24 (60.0) | 44 (52.4) |  |  |
| Supervisor<br>nurse<br>(Zhuguan<br>hushi) | 5 (11.4) | 15 (37.5) | 20 (23.8) |  |  |
| Education, n<br>(%) |  |  |  | 2.161 (2) | 0.339§ |
| Associate<br>degree | 6 (13.6) | 2 (5.0) | 8 (9.5) |  |  |
| Bachelor's<br>degree | 37 (84.1) | 36 (90.0) | 73 (86.9) |  |  |
| Master's<br>degree | 1 (2.3) | 2 (5.0) | 3 (3.6) |  |  |
| Handovers<br>per month, n<br>(%) |  |  |  | 0.033 (2) | 0.984 |
| ≤5 | 13 (29.5) | 12 (30.0) | 25 (29.8) |  |  |

|  |  |  |  |
| --- | --- | --- | --- |
| 6-10 | 26 (59.1) | 23 (57.5) | 49 (58.3) |
| 11-20 | 5 (11.4) | 5 (12.5) | 10 (11.9) |

† Two cells (25.0%) have expected count <5 (minimum 3.33).
‡ Fisher's exact test $p = 0.006$ (2-tailed).
§ Four cells (66.7%) have expected count <5 (minimum 1.43); result should be
interpreted with caution

### Known-groups validity

Discriminant validity was assessed by comparing handover scores among nurses with different levels of clinical experience.

The results showed that nurses with more clinical experience had significantly higher total scores than those with less experience (Z = −4.772, p < 0.001).

At the dimensional level, statistically significant differences were observed in the “Identification” (Z = −2.241, p = 0.025), “Context” (Z = −3.593, p < 0.001), and “Assessment” (Z = −3.628, p < 0.001) dimensions. In contrast, no statistically significant differences were found in the “Situation” (Z = −1.451, p = 0.147) and “Recommendation” (Z = −1.785, p = 0.074) dimensions (Table 3).

**Table 3.**
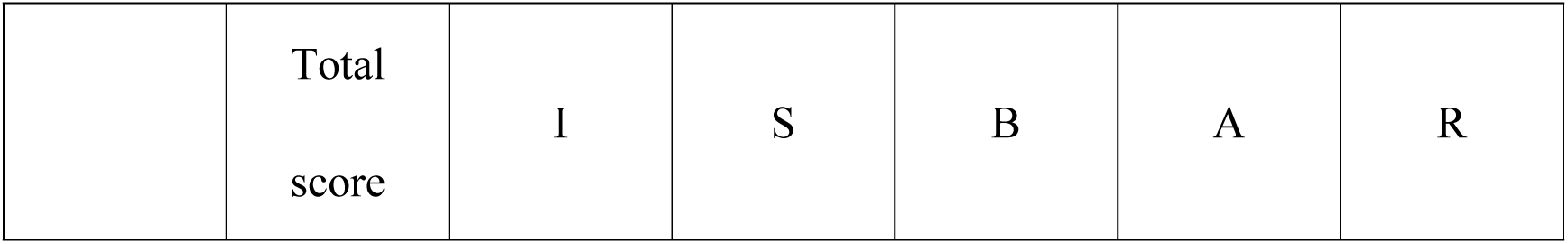
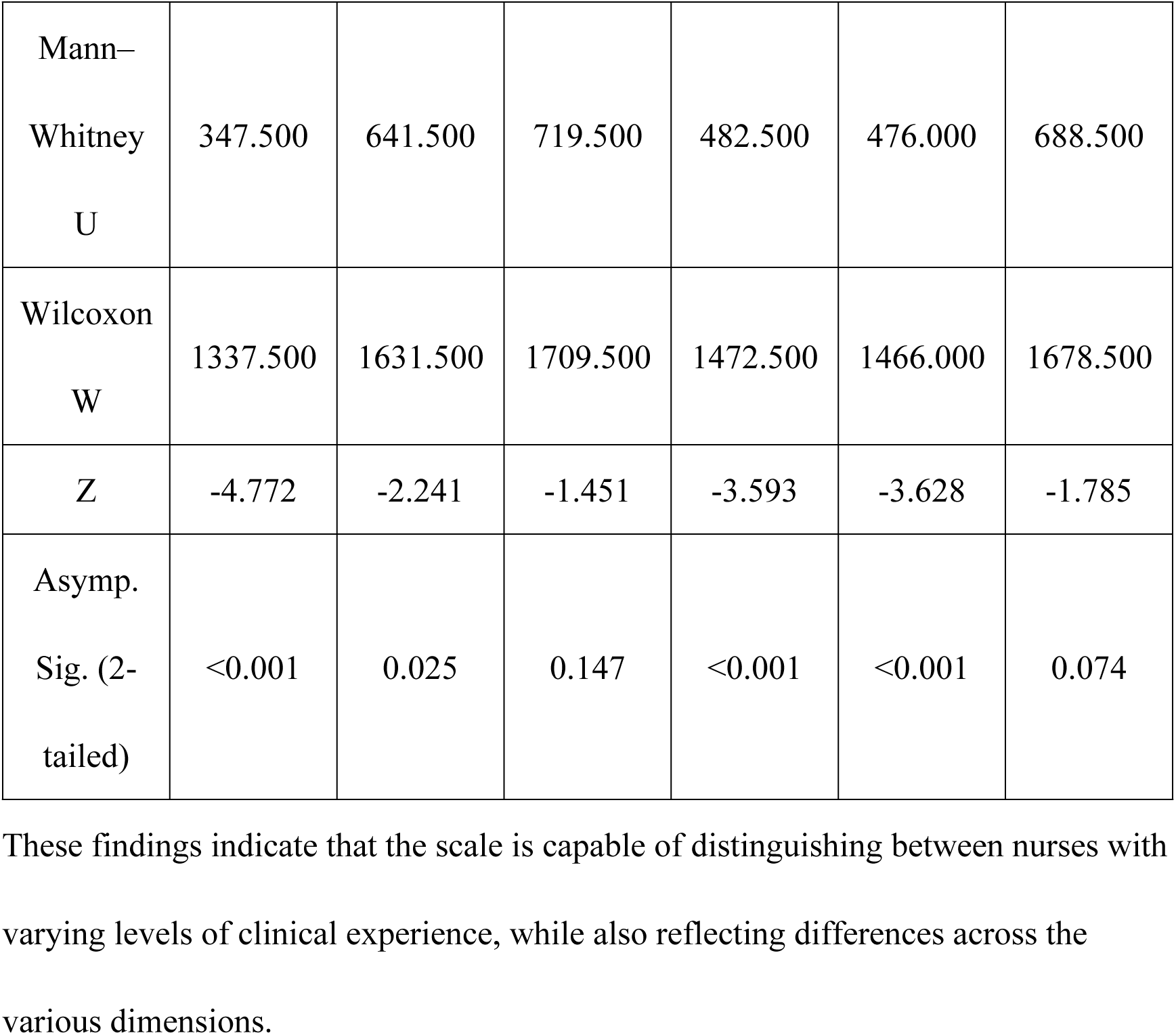
Discriminant validity. These findings indicate that the scale is capable of distinguishing between nurses with varying levels of clinical experience, while also reflecting differences across the various dimensions.

## Discussion

This study conducted a cross-cultural adaptation and psychometric validation of the ISBAR Structured Handover Observation Tool. The results indicate that the Chinese version of the scale demonstrates good content validity (S-CVI/Ave = 0.99) and inter-rater reliability (total score ICC = 0.885, 95% CI: 0.795–0.935) and is effective in distinguishing nurses with different levels of clinical experience (Z = −4.772, p < 0.001). These findings support the tool’s applicability in assessing the completeness of information transfer during patient handoffs from the ICU to general wards.

In terms of content validity, the S-CVI/Ave for this scale was 0.99, which is higher than the S-CVI value (0.91) reported by Tangvik et al. [1] in the original study. This indicates that the Chinese version maintains good content validity following cross-cultural adaptation and demonstrates high inter-rater reliability. Regarding inter-rater reliability, the original study did not report this metric; however, this study found that the ICC for the total score was 0.885 (95% CI: 0.795–0.935), with ICCs for each dimension ranging from 0.789 to 0.941, providing new evidence of the instrument’s reliability. Furthermore, the discriminant validity results of this study showed that nurses with higher seniority scored significantly higher than those with lower seniority (Z = −4.772, p < 0.001), which is consistent with previous research: He et al. [18] found that years of service were a significant predictor of nurses’ clinical communication skills (F = 56.212, p < 0.001), while Qiu et al. [24] also reported a positive correlation between years of experience and medical narrative competence. These findings collectively support the positive correlation between clinical experience and structured communication performance and provide external evidence for the construct validity of the ISBAR observation tool.

However, in terms of discriminant validity, not all dimensions showed statistically significant differences. The “Situation” and “Recommendation” dimensions were not statistically significant, which may reflect their lower sensitivity to differences in clinical experience. A few possible reasons for this result persist. First, the information in the “Situation” dimension is relatively standardized and procedural; regardless of seniority, nurses are required to clearly explain the patient’s current condition and subsequent treatment plan during handoffs, resulting in minimal performance differences among nurses of varying experience levels on this dimension [25]. Second, the “Recommendation” dimension contains only two items; the small number of items may limit its discriminatory power. Third, this scale uses a dichotomous (“yes/no”) scoring method, focusing on assessing the completeness of information transmission [26], while failing to capture the deeper advantages of senior nurses in terms of clinical insight on the “Situation” dimension and decision-making quality on the “Recommendation” dimension.

This study has several limitations. First, although dichotomous scoring effectively assesses whether information was conveyed, it does not reflect the quality or depth of that transmission. Previous studies have noted that while dichotomous checklists can distinguish between operators of different skill levels, they cannot provide detailed feedback [26]. Second, the “Recommendations” dimension includes only two items, which may limit its statistical discriminatory power. Third, the sample in this study was drawn from two tertiary hospitals, and the generalizability of the results to primary care settings or other cultural contexts requires further validation. Fourth, despite informing nurses in advance that video recordings were for research purposes only and would not affect performance evaluations, the recording process may still have induced a Hawthorne effect – a phenomenon in which individuals alter their behavior due to awareness of being observed [27]. This may have led to transiently higher handover scores than would occur in routine, unobserved practice. In summary, the Chinese version of the ISBAR Structured Handover Observation Tool demonstrates good reliability and validity in assessing patient handover from the ICU to general wards.

## Conclusions

This study successfully adapted the ISBAR Structured Handover Observation Tool for a cross-cultural context and validated the reliability and validity of its Chinese version in assessing patient handover from the ICU to general wards. The Chinese version of the scale demonstrated good content validity, inter-rater reliability, and discriminant validity, enabling an objective assessment of adherence to the ISBAR framework during the handover process. This tool provides clinicians with a standardized assessment method that can be used for quality monitoring, continuous feedback, and targeted training interventions, thereby helping to bridge the gap between recommended communication frameworks and clinical practice. Future research should develop more refined evaluation tools (such as those using a rating scale) and further validate their applicability across multicenter settings and healthcare facilities at various levels.

## Data Availability

Due to ethical restrictions and participant confidentiality agreements approved by the Medical Ethics Committee of the University of South China (Approval No. 2025-0148), the raw data supporting the findings of this study cannot be made publicly available. De-identified data may be obtained from the corresponding author upon reasonable request, subject to institutional approval and data use agreements.

## Acknowledgments

The authors would like to thank all the nurses who participated in this study for their valuable contributions. We would also like to express our gratitude to the experts who were involved in the translation and cross-cultural adaptation process of this instrument. In addition, we acknowledge the support provided by the participating hospitals. We would like to thank Editage (www.editage.cn) for English language editing.

## Author contributions

**Conceptualization:** Na Ni; Yuting Wang; Qi Wang; Jiajia Ding.

**Data curation**: Na Ni; Jiajia Ding; Tong Liu.

**Formal analysis**: Na Ni; Yuting Wang.

**Funding acquisition**: Bixia Zhao.

**Investigation**: Na Ni; Yuting Wang; Qi Wang; Jiajia Ding; Tong Liu.

**Methodology**: Na Ni; Yuting Wang; Qi Wang; Jiajia Ding; Tong Liu.

**Project administration**: Na Ni; Bixia Zhao.

**Resources**: Na Ni; Yuting Wang; Qi Wang.

**Supervision**: Na Ni; Bixia Zhao.

**Writing – original draft**: Na Ni; Yuting Wang; Qi Wang; Bixia Zhao.

**Writing – review & editing**: Na Ni; Yuting Wang; Qi Wang; Jiajia Ding; Tong Liu; Bixia Zhao.

## Supporting information

S1 Dataset. Raw data of ISBAR observation scores and nurse characteristics. (XLSX)

This file contains the raw scores of the ISBAR observation tool for 233 handover cases and the baseline characteristics (age, professional title, education, etc.) of the 84 participating nurses.

S2 Dataset. Expert consultation records for content validity evaluation. (XLSX)

This file contains the detailed I-CVI ratings from the expert panel across two rounds of consultation.

S3 Dataset. Translation and cross-cultural adaptation records. (XLSX)

This file documents the forward translation, back-translation, and expert review process for the Chinese version of the ISBAR observation tool

